# From Bimodal to Unimodal: The Transformed Incidence of Osteosarcoma in the United States

**DOI:** 10.1101/2023.09.04.23294332

**Authors:** Emma Kar, Amrit Ammanamanchi, Miranda Yousif, Saroja Devi Geetha, Kendall Schwartz, Arya Mishra, Jiali Ling, Kristie Nneoma Nonyelu, Bijun Sai Kannadath

## Abstract

**Background:** Osteosarcoma is the most common primary bone malignancy. It has classically been described as having a bimodal incidence by age, with the first peak in the second decade of life and the second peak after 65 years of age. We sought to identify whether the bimodal incidence distribution still exists for osteosarcoma using the National Cancer Institute’s Surveillance, Epidemiology, and End Results (SEER) and the National Inpatient Sample (NIS) (Healthcare Cost and Utilization Project (HCUP), Agency for Healthcare Research and Quality) databases.

**Methods:** Incidence rates of osteosarcoma between 1975-2020 were analyzed by age, year of occurrence, sex, race, and primary tumor site using the SEER program data. The age of maximum incidence in patients 40 years of age and older was analyzed by calendar year to observe for a consistent second peak. Fisher’s Exact test was also conducted with the SEER data to assess if there was a significant association between the occurrence of osteosarcoma in patients 35-64 years old and those 65+ years old. Incidence of tumors of the long bones of the lower limbs using ICD-9 and 10 diagnosis codes from the NIS discharge database 2012-2019 were also analyzed for comparison.

**Results:** A total of 1,779 cases of osteosarcoma were reported in the SEER database from 1975-2020. Across the 46 calendar year span, a consistent first peak appeared in the second decade of life alternating between the 10-14 and 15-19 age groups. In the 40+ cohort, the age of the highest incidence varied widely each calendar year: being found in age groups 40-44, 45-49, 50-54, 55-59, 60-64, 65-69, 70-74, 75-79, & 85+ years old (5, 3, 5, 1, 2, 1, 2, 1, & 1 times respectively). For 25 years of the 46-year span, the max was shared by more than one age group. Overall, no single age group from the 40+ age group held a consistent maximum from 1975-2020.

Using the NIS program and similar methodology, we analyzed 86,100 discharges of lower limb long bone tumors from 2012-2019 as a reference. The NIS data exhibited nearly identical patterns, with a unimodal incidence in adolescence and no obvious second peak.

**Conclusion:** Our analysis shows that the incidence of osteosarcoma is no longer bimodally distributed. This may be due to the increased use of bisphosphonates in recent years for the effective management of Paget’s disease, leading to a decline in late-age bone tumors. Our findings suggest the need to update our understanding of the epidemiology of osteosarcoma, as it should be considered more as a disease of adolescence with only sporadic incidence later on in older age.

## Introduction

The most common primary tumor of the bones is osteosarcoma.^1^ It is conventionally described as having a bimodal incidence by age, with the first mode in the teenage years and the second mode after 65 years of age.^2-7^

We sought to examine the current age distribution for the incidence of osteosarcoma. We used two large United States specific health databases: The National Cancer Institute’s Surveillance, Epidemiology, and End Results (SEER) program which collects data on cancer incidence from various cancer registries, serving about 47.9 percent of the U.S. Population, and The National Inpatient Sample (NIS) dataset (Healthcare Cost and Utilization Project (HCUP), Agency for Healthcare Research and Quality), which is the largest publicly available database of inpatient discharge data in the country.^8,9^

## Materials and Methods

### SEER

We extracted data from the SEER Research Data, 8 Registries, Nov 2021 Sub (1975-2020).^8^ SEER*Stat 8.4.1 was utilized to extract frequency and incidence rates. Data analysis was done using Python via Jupyter notebooks.^10^ Osteosarcoma cases were classified by site morphology (4.1 Osteosarcoma) according to adolescents and young adults (AYA) site recode 2020 Revision and primary site (C40.2 - Long bones of lower limb and associated joints) according to the World Health Organization (WHO) Classification of Tumors International Classification of Disease for Oncology, 3rd edition (ICD-O-3 codes). The incidence of osteosarcoma of the long bones of the lower limbs was analyzed for trends over calendar years across each age group (in four-year wide bins from 1-4 years to 80-84 years, and 85+ years as provided by SEER).

We investigated the presence of a second incidence peak by analyzing the incidence in patients 40 years and older, looking for the peak age of incidence in each calendar year. The number of times each age group had a unique maximum was tallied and assessed for trends of a ‘true’ second incidence peak. Any calendar year where more than one age group held the maximum was excluded from the final tally.

We conducted a Fisher’s exact test for each calendar year in the SEER data, assessing if there was a significant difference between the occurrence of osteosarcoma in patients 35-64 years old versus those 65+ years old (the age group classically described as constituting the second peak).^3-7^ The p-values and odds ratio for each test across the 46 calendar year period were assessed for chronological trends over time. Statsmodel Version 0.13.5 in Python was used to conduct the Fisher’s exact test with the significance level set at 0.05. Finally, we assessed for trends in incidence by biological sex across the different age groups in the SEER data.

### NIS

We also collected data from the NIS for 2012-2019.^9^ Since tumors are not classified by histological subtype within the NIS database, we used ICD-9 and ICD-10 codes which report the primary tumor location. Patient data were extracted for primary tumors of the long bones of the lower limbs (ICD-9: 17.07; ICD-10: C40.20, C40.21, and C40.22) from NIS. The incidence and incidence rates of primary tumors of the long bones of the lower limbs were assessed by age for trends from year to year over 2012-2019. The same assessment analyzing the incidence in patients 40 years and older was done on the NIS dataset, searching for the peak age of incidence in each calendar year. Finally, we assessed for trends in incidence by biological sex across the different age groups in the NIS data.

## Results

### SEER

There were 1,779 (790 females and 989 males) cases of osteosarcoma of the long bones of the lower limbs reported in the SEER 8 database from 1975-2020. Of them, 1,440 cases were reported in patients less than 40 years of age and 339 in patients 40 years or older. There was a consistent first incidence peak alternating between the age groups of 10-14 and 15-19. (Figures 1, 2, & 3).

**Figure 1:**
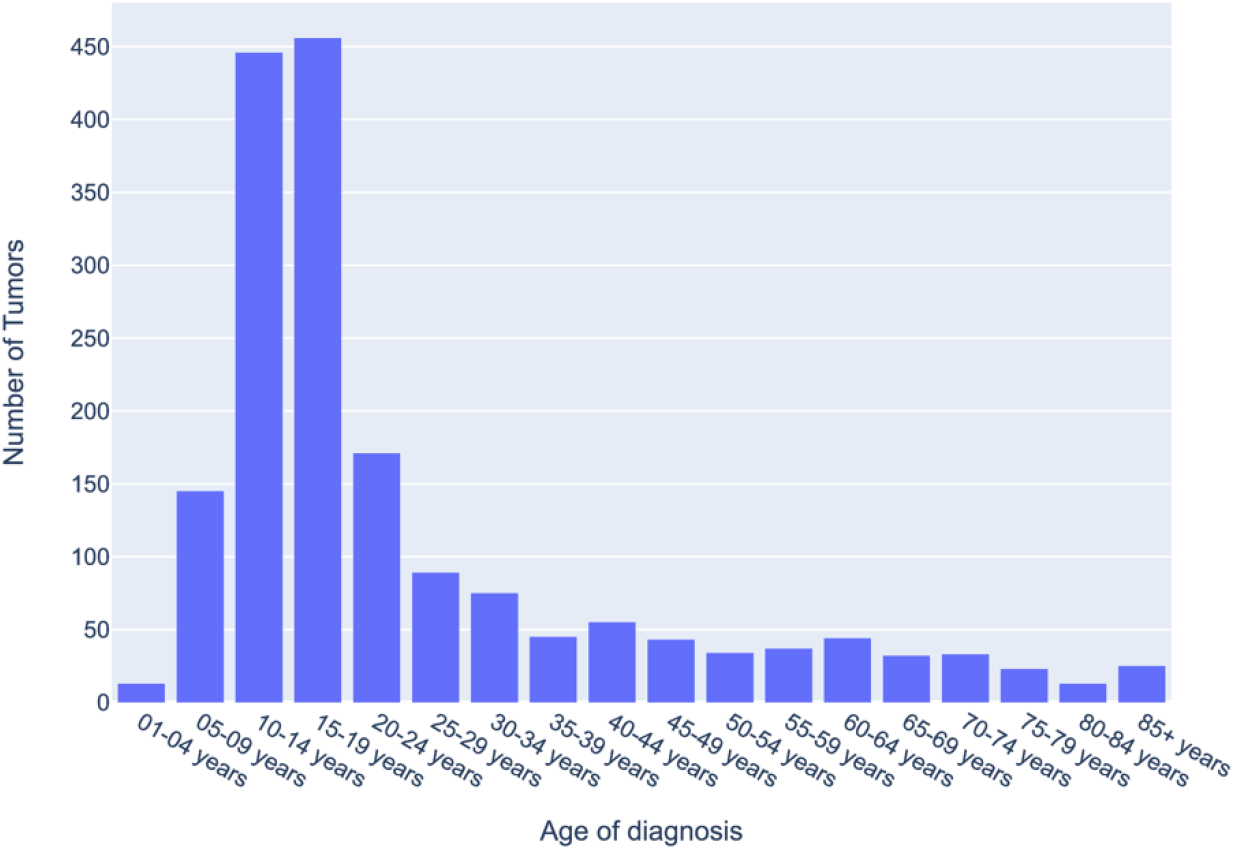
Distribution of Osteosarcoma of long bones of the lower limbs based on age at diagnosis from SEER data 1975-2020: Total incidence of Osteosarcoma of the long bones of the lower limbs based on Surveillance, Epidemiology, and End Results (SEER) 8 database, 1975-2020, by age at initial diagnosis. The number of tumors was totaled in each age group across the 46-year span.

**Figure 2:**
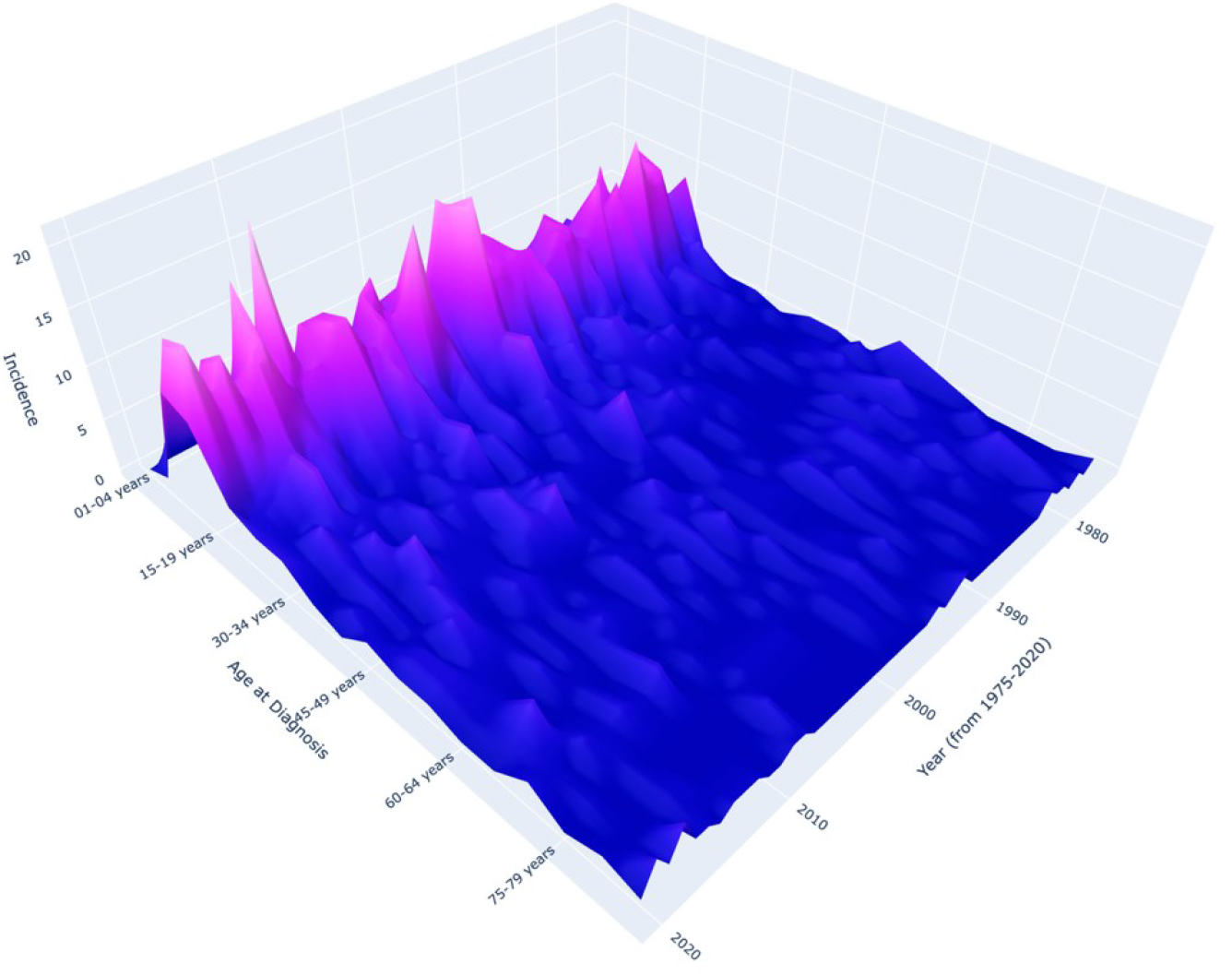
Incidence of Osteosarcoma of Long Bones of Lower Limbs from SEER data 1975-2020: 3D Representation of the Incidence of Osteosarcoma of the long bones of the lower limbs based on Surveillance, Epidemiology, and End Results (SEER) 8 database, 1975-2020, by age at initial diagnosis and year of diagnosis. The change in color to a lighter purple corresponds to an increased number of tumors reported in that age group that particular calendar year.

**Figure 3:**
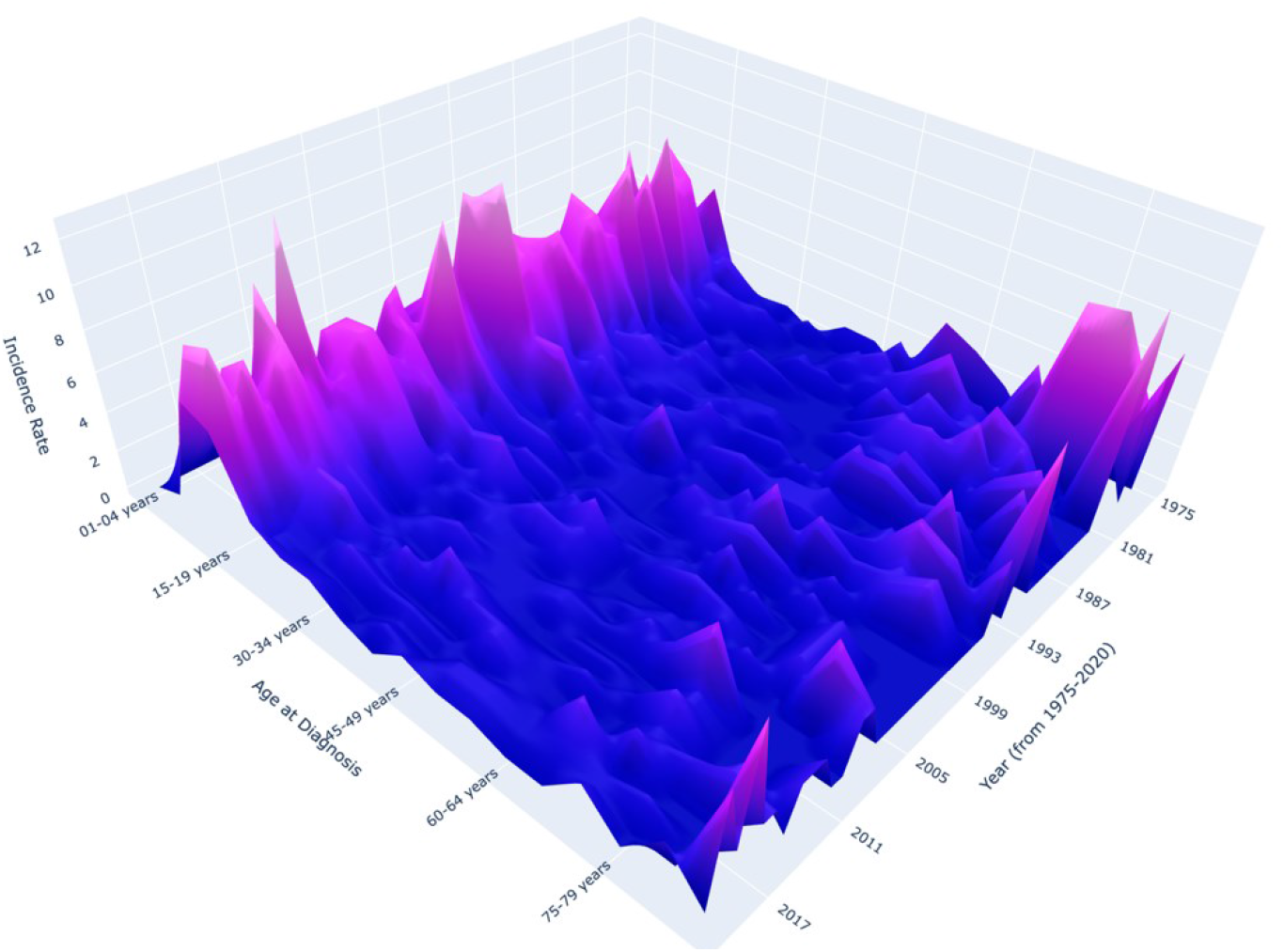
Incidence Rate of Osteosarcoma of Long Bones of Lower Limbs from SEER data 1975-2020: 3D Representation of the Incidence Rate of Osteosarcoma of the long bones of the lower limbs based on Surveillance, Epidemiology, and End Results (SEER) 8 database, 1975-2020, by age at initial diagnosis and year of diagnosis. The change in color to a lighter purple corresponds to an increased incidence rate of osteosarcoma reported in that age group that particular calendar year.

In patients 40 years and older, there was no single age group that held a consistent peak incidence in osteosarcoma each calendar year across the 46-year period. (Figure 1 & 2). The location of the maximum incidence varied each calendar year within this sub-cohort. In the sub-cohort analysis, there were 21 years out of the 46 calendar years where there was only a single maximum in the 40+ age group (Figure 4). The 40-44 years old and 50-54 years old age groups both had 5 unique maximums each, with the other age groups holding fewer (Table 1).

**Table 1:** Number of Unique Maximums of Osteosarcoma of the Long Bones of the Lower Limbs for 40+ years old in SEER data 1975-2020: There were 21 out of 46 years where there was a unique maximum in those 40 years and older based on Surveillance, Epidemiology, and End Results (SEER) 8 database across each calendar year from 1975-2020. The total number of times each age group held a unique maximum was tallied above.

| Age group (years) | Number of unique maximums |
| --- | --- |
| 40-44 | 5 |
| 45-49 | 3 |
| 50-54 | 5 |
| 55-59 | 1 |
| 60-64 | 2 |
| 65-69 | 1 |
| 70-74 | 2 |
| 75-79 | 1 |
| 85+ | 1 |

**Figure 4:**
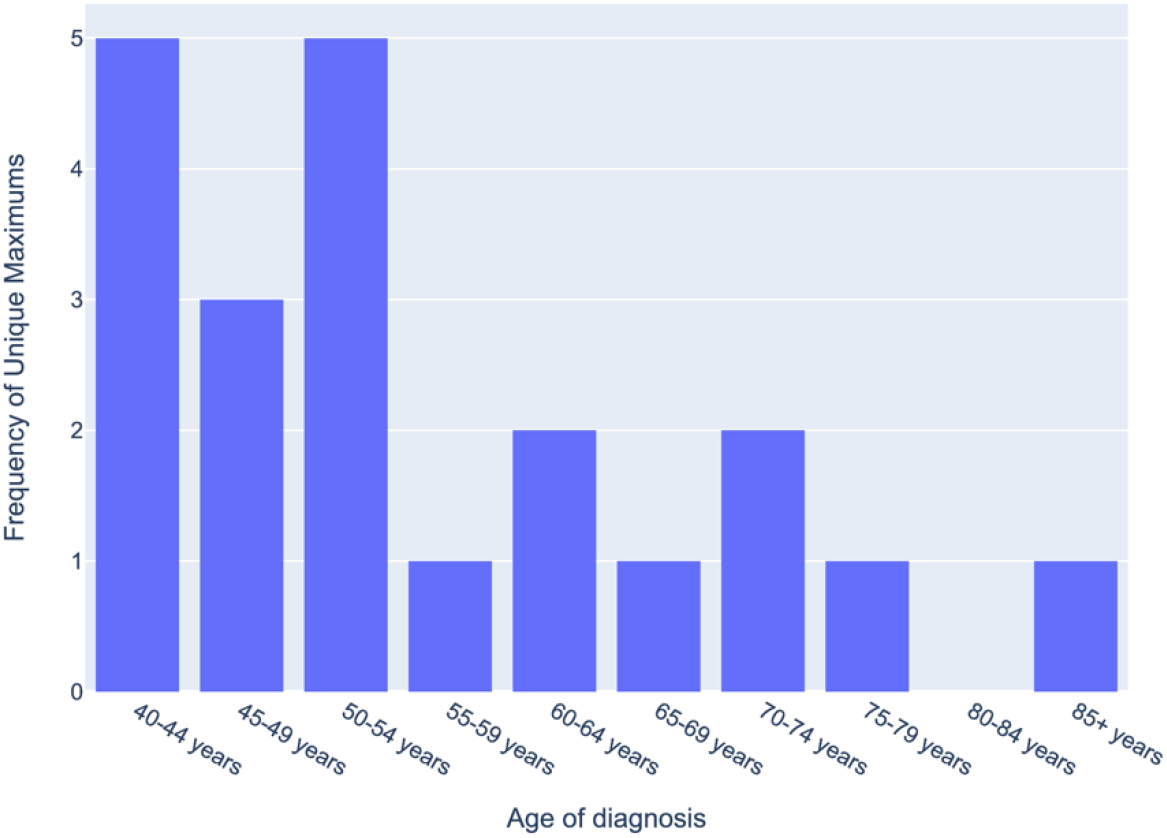
Frequency of Unique Maximums of Osteosarcoma of the Long Bones of the Lower Limbs for 40+ years old in SEER data 1975-2020: Sub-cohort analysis assessing for frequency of unique maximums of Osteosarcoma of the long bones of the lower limbs in those 40 years and older based on Surveillance, Epidemiology, and End Results (SEER) 8 database across each calendar year from 1975-2020.

The results of Fisher’s Exact test revealed a significant difference (p-value<0.05) between the two groups during 2 calendar years: 1997 and 2017. There was no significant difference (p-value>0.05) found between the two groups during the remaining 44 calendar years. Overall, the resulting p-values tended to increase as the years progressed to be greater than 0.05, displaying a chronological trend in decreasing significance (Table 2). The odds ratio of incidence between the two cohorts also showed a decline over the years, trending closer to 1 as the calendar years progressed (Figure 5).

**Table 2:** P-value Results from Fisher’s Test Comparing Incidence in Age Groups 35-64 years vs 65+ years in SEER data 1975-2020: P-value results from Fisher’s Test for the SEER data each calendar year (1975-2020), assessing for a significance in the incidence of osteosarcoma between age groups 35-64 years and 65+ years. All significant p-values (p<0.05) are in red, indicating those years where a significant difference in the incidence of osteosarcoma between the two age groups (35-64 and 65+) exists.

| Year | p-value | Year | p-value | Year | p-value | Year | p-value |
| --- | --- | --- | --- | --- | --- | --- | --- |
| 1975 | 1.00000 | 1985 | 0.11156 | 1995 | 0.07118 | 2005 | 0.74884 |
| 1976 | 1.00000 | 1986 | 0.18201 | 1996 | 0.46835 | 2006 | 0.62243 |
| 1977 | 0.10105 | 1987 | 1.00000 | 1997 | 0.04001 | 2007 | 0.53366 |
| 1978 | 0.37664 | 1988 | 0.07672 | 1998 | 1.00000 | 2008 | 0.69168 |
| 1979 | 0.05687 | 1989 | 0.33813 | 1999 | 0.69083 | 2009 | 0.34727 |
| 1980 | 0.07395 | 1990 | 0.48703 | 2000 | 1.00000 | 2010 | 0.16762 |
| 1981 | 0.12229 | 1991 | 0.64644 | 2001 | 0.53978 | 2011 | 1.00000 |
| 1982 | 0.17918 | 1992 | 0.42175 | 2002 | 1.00000 | 2012 | 0.68250 |
| 1983 | 0.60844 | 1993 | 1.00000 | 2003 | 0.62348 | 2013 | 0.07204 |
| 1984 | 0.38349 | 1994 | 1.00000 | 2004 | 1.00000 | 2014 | 0.54095 |

**Figure 5:**
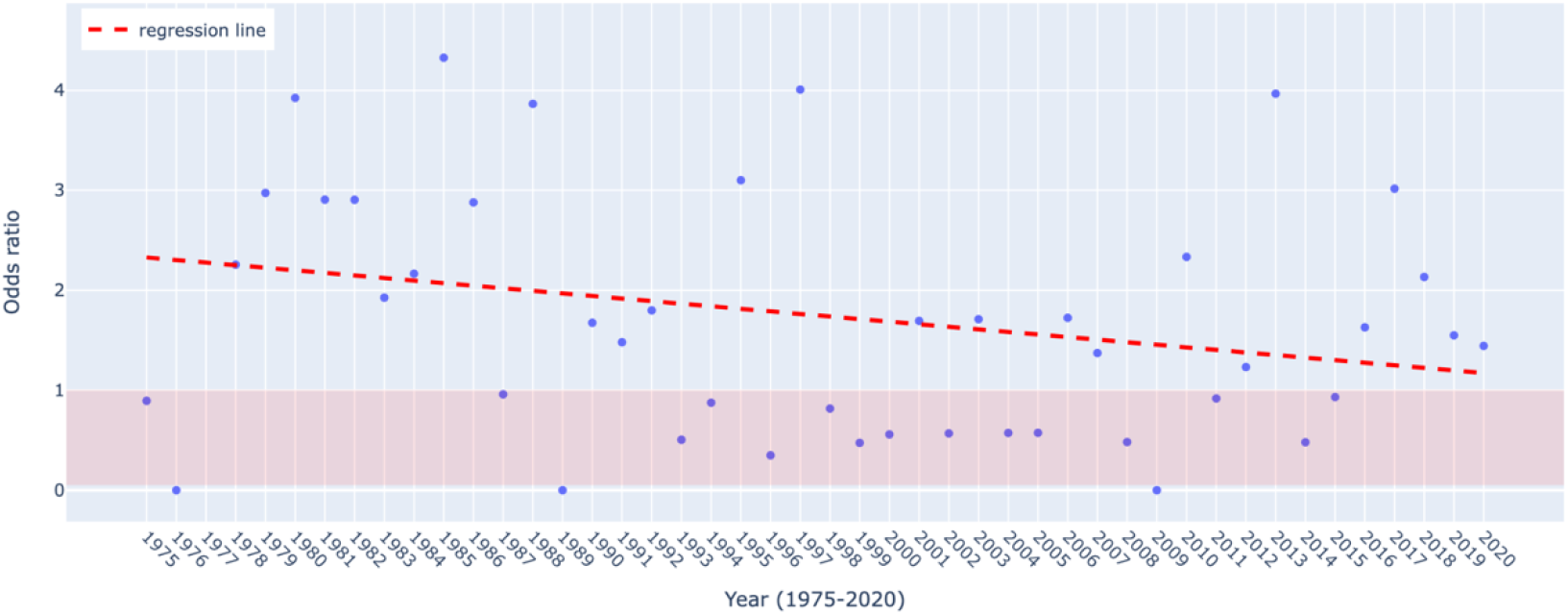
Odds Ratio from Fisher’s Test Across the SEER data 1975-2020 between 35-64 years & 65+ years: Odds ratio results from Fischer’s Exact Test for the SEER data each calendar year (1975-2020), assessing for a significance in the incidence of osteosarcoma between age groups 35-64 years and 65+ years. The red line represents the best fit regression line across the 46 year period, demonstrating that the odds ratio decreases over the years due to the negative slope of the line.

### NIS

There were 86,100 discharges of tumors of the long bones of the lower limbs reported in the NIS data from 2012-2019 (Figure 6), which showed close to identical trends, displaying only one peak of incidence and incidence rates in early adolescence, peaking around 10-15 years.

**Figure 6:**
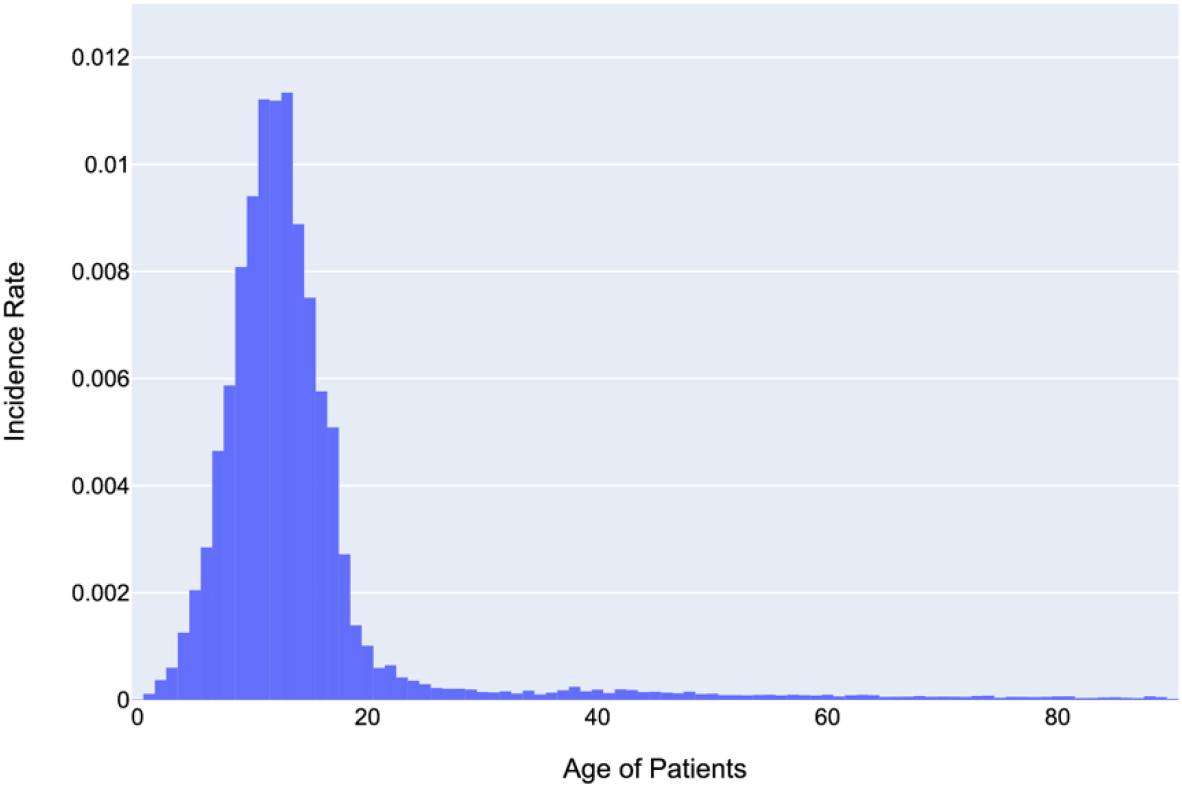
Age of Patients with Tumors of Long Bone of the Lower Limbs (NIS data 2012-2019): Incidence rate of tumors of the long bones of the lower limbs based on the National Inpatient Sample (NIS) from 2012 to 2019 by age at diagnosis. The incidence rate was calculated based on the total number of tumors in each age group across the 8-year span of data.

There was a unique maximum in the 40+ years old sub-cohort each year from 2012-2019. The subgroup that held the maximum, varied between years: 3 times in 42 years, 3 times in 43 years, 1 time in 45 years, and 1 time in 48 years (Figure 7).

**Figure 7:**
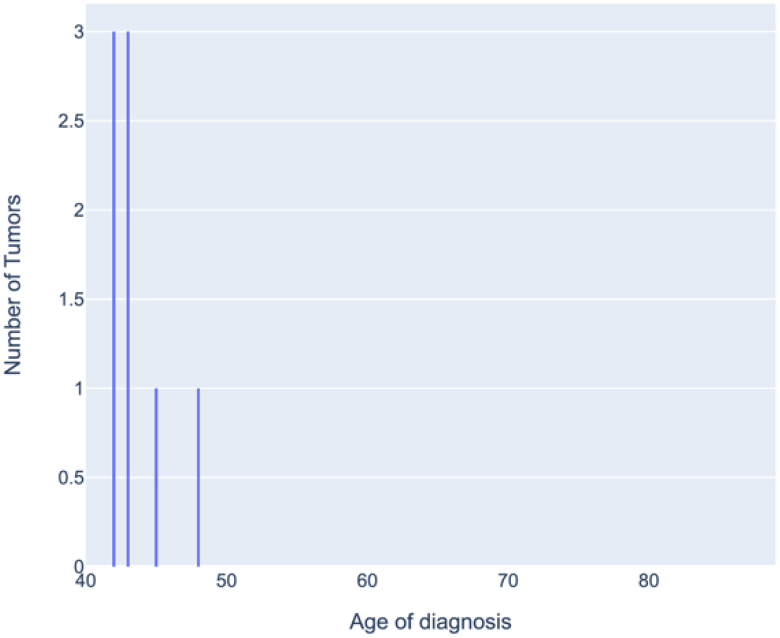
Frequency of Max Incidence Rate of Tumors of the Long Bones of the Lower Limbs for 40+ years old in NIS data 2012-2019: Sub-cohort analysis of the 40+ years old age group assessing the frequency of maximum incidence rate of tumors of the long bones of the lower limbs in the National Inpatient Sample (NIS) from 2012 to 2019. Each time an age group held the maximum incidence rate for one calendar year was tallied and results are shown here.

## Discussion

This bimodal incidence was first proposed in a seminal single-author study published in the British Journal of Cancer in 1955. In this study, researcher C.H. Price independently analyzed 67 cases of osteogenic sarcoma, graphing their incidence.^11^ Price observed two peaks in incidence, first in the 2nd decade of life and then around the 7th and 8th decades.^11^ Price followed up this work with a monograph postulating how the drastic increase in bone tumors in adolescents correlates to bone activity, since there is rapid growth and increased activity occurring at the metaphyseal plates during puberty.^12^ These two studies have been pivotal in shaping the current understanding of the incidence of osteosarcoma.

### Primary Incidence Peak

When assessing the overall incidence of osteosarcoma by age, we found a significant peak across all calendar years in both the SEER and NIS data sets that suggest a spike in its incidence around the onset of puberty. This is attributable to an increase in activity at the metaphyseal plates leading to the rapid increase in growth that occurs around the time of puberty.^2,6,12^ Furthermore, the most common sites for osteosarcoma are the distal femur, proximal tibia, and proximal humerus, which are all long bones that grow significantly during puberty.^4,6^ This further reinforces the idea that osteosarcoma may be associated with increased activity at the metaphysis of bones, supporting the original theories proposed by Price.^12,13^

### Second Incidence Peak

Overall, the SEER 8 database from 1975-2020 lacks a second incidence peak displaying no bimodal age distribution of osteosarcoma. In contrast to the strong spike of the first peak seen in 10-19 year-olds, there is no age group in the 40+ years old sub-cohort that can indicate a second peak. However, assessing trends only by incidence does not account for the change in population size each calendar year within each age group.

Since the size of the population under 40 years old is much greater than the population that is 40 years and older, the occurrence of a second incidence peak could be ‘hidden.’ The 3D model of the incidence rate of osteosarcoma (Figure 3) is a representation of the incidence adjusted for the population in each age group each calendar year. This model also displays a strong first incidence peak consistently over all 46 calendar years, however, a consistent second peak is absent.

Our further analysis which assessed for a single unique maximum within the 40+ years old group also further reinforced the lack of a strong second incidence peak, as two age groups (40-44 years and 50-54 years) “won” the tally for this assessment. Likewise, these two age groups are significantly younger than the proposed site for a second incidence peak (65+ years old). Furthermore, since these results (Figure 4) showed no normalized curve or tapering edges around one particular age group, this assessment supported the lack of a second incidence peak in the SEER data. The declining odds ratio across the calendar years further confirms that the SEER osteosarcoma data is not bimodally distributed by age (Figure 5).

### Comparison to NIS Data

The NIS data clearly displayed a flattening of the incidence rate in the 40+ age groups thereby reinforcing a lack of a second peak. (Figure 6). No particular age group demonstrated a unique maximum incidence rate. However, this analysis only accounts for tumors of the long bone of the lower limbs which are not entirely osteosarcoma, though osteosarcoma overall is the most common bone tumor. For this reason, the NIS results are supportive of the SEER results but are not conclusive on its own.

### Gender Distribution

Our analysis using SEER 8 and NIS database revealed comparable findings with previous studies in terms of incidence rates by biological sex, though we observed an overall higher incidence of osteosarcoma in males when compared to females (Figures 8 & 9).^7^

**Figure 8:**
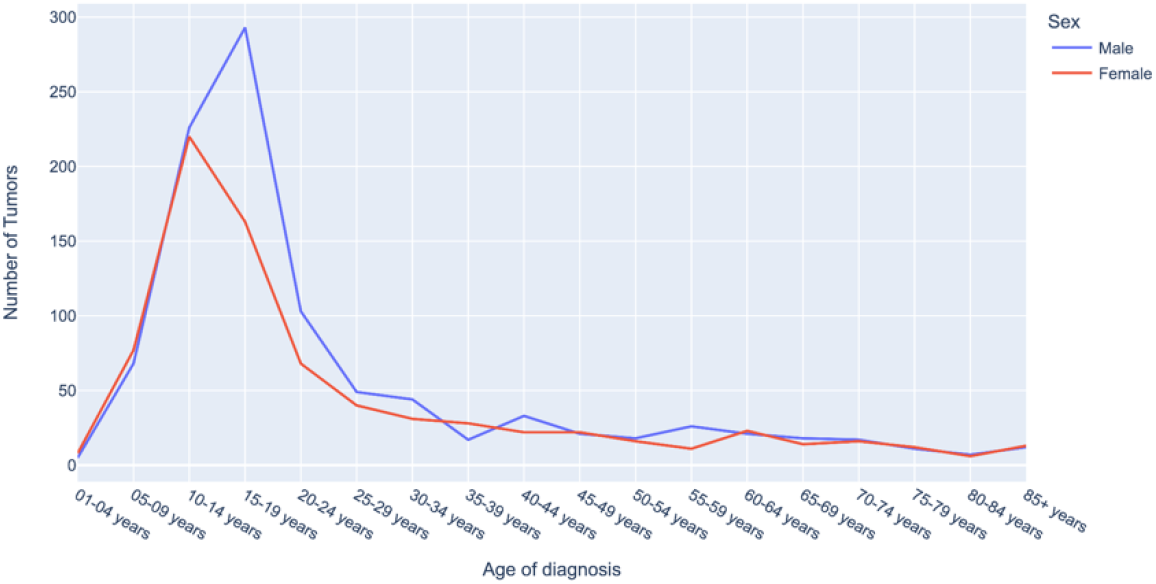
Sex distribution of Incidence of Osteosarcoma of the Long Bones of the Lower Limbs by Age for SEER data 1975-2020: Total incidence of Osteosarcoma of the long bones of the lower limbs based on Surveillance, Epidemiology, and End Results (SEER) 8 database, 1975-2020, by age at initial diagnosis and sex. The number of tumors was totaled in each age group across the 46-year span.

**Figure 9:**
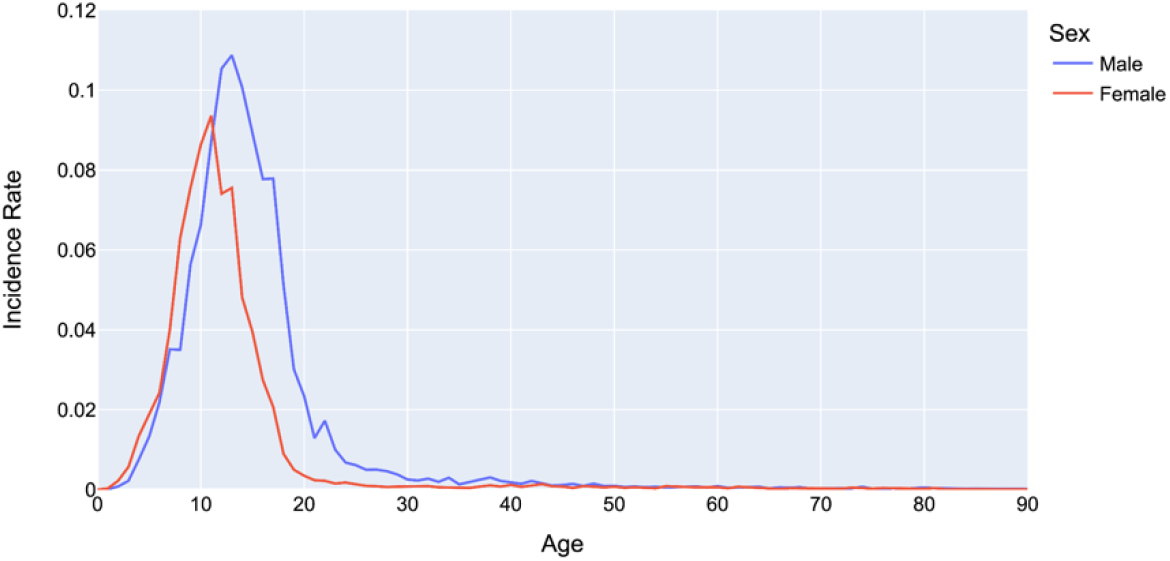
Sex distribution of Incidence Rate of Osteosarcoma of the Long Bones of the Lower Limbs by Age for NIS data 2012-2019: Incidence rate of tumors of the long bones of the lower limbs based on the National Inpatient Sample (NIS) from 2012 to 2019 by age at diagnosis and sex. The incidence rate was calculated based on the total number of tumors in each age group across the 8-year span of data.

Historically, the occurrence of a second peak was attributed to secondary malignancy arising from pre-existing Paget’s disease.^2,5^ The pathogenesis of Paget’s disease begins with overactive osteoclasts, promoting excessive bone resorption, and later triggering higher activity of osteoblasts, the precursor cells to osteosarcoma.^15^ Today, there are highly effective pharmacotherapeutic measures for Paget’s disease that specifically target reducing osteoclast function, like oral bisphosphonates and calcitonin injections, which can result in near complete control of the disease.^15-17^ In particular, with the introduction of bisphosphonates around the 1970s for the treatment of Paget’s disease, there has been a noticeable decrease in the prevalence of osteosarcoma arising from this disease.^16,18^ Evidence for this can be found in the Japanese population, where the development of Paget’s disease is rare and a second peak has never been observed.^2,14^ Thus the change from bimodal to unimodal can be attributed to the advancements in the management of Paget’s Disease in the West.

Overall, these findings offer a significant shift in our understanding of the epidemiological spread of osteosarcoma. Understanding the change in incidence might help us to better approach spreading awareness and diagnosing osteosarcoma and likewise focus on the most affected age groups. Tracking the trends in age groups and genders affected can also help to increase our insight into potential risk factors associated with this rare neoplasm, such as environmental exposures, lifestyle measures, or other genetic associations. Developing this updated understanding of the changing age distribution for osteosarcoma occurrence can also aid in designing age-specific screening strategies and also determining the limits for diagnostic tools that vary based on age and gender. While this study only investigated the occurrence of primary osteosarcoma tumors (not recurrence), there might also be a need to look at the changes in trends of recurrence of osteosarcoma as treatments have significantly changed. Prior to 1970, one of the only treatments for osteosarcoma was amputation and about 80% of individuals died from metastases to the lungs, with a 5-year survival rate of 20%.^4,5^ In more recent times, the treatment of choice for osteosarcoma is neoadjuvant chemotherapy, followed by limb-salvage surgery and chemotherapy.^19,20^ Treatment can also include the use of newer technology like radiation.^19,20^ Continuing to follow these trends in osteosarcoma will help investigate the impact of current treatment protocols and possibly offer suggestions for improvements in these therapies that might better treat the age groups affected.

## Conclusion

Our analysis of two large US databases points towards a Unimodal pattern of incidence of Osteosarcoma. This is a significant shift from existing understanding of the bimodal incidence. In other areas, this study supports previous findings that osteosarcoma occurs more frequently in men than in women. Our findings suggest that literature must be updated to better inform healthcare providers of the true epidemiology and to focus diagnostic effort in the teenage years while screening for osteosarcoma.

## Data Availability

All data produced in the present study are available upon reasonable request to the authors.

